# Correlation between hypophosphatemia and the severity of Corona Virus Disease 2019 patients

**DOI:** 10.1101/2020.03.27.20040816

**Authors:** Xiaobo Xue, Jing Ma, Yuxia Zhao, Aibin Zhao, Xiaohong Liu, Wei Guo, Fang Yan, Zhixin Wang, Yanqing Guo, Mengbai Fan

## Abstract

**Objective:** Retrospectively analyze the clinical data of Corona Virus Disease 2019 (COVID-19) patients and explore the value of serum phosphorus level in evaluating the severity and prognosis of the disease.

**Methods:** COVID-19 patients transferred from the first emergency ward of Taiyuan fourth people’s Hospital from February 8 to March 3, 2020 were enrolled. The information of general conditions, clinical manifestations, laboratory indexes, nucleic acid detection and treatment were collected. The changes of blood phosphorus level and absolute value of lymphocytes in ordinary and severe/critical patients were recorded and compared.

**Results:** A total of 32 patients with COVID-19 were collected, including 12 cases of common type and 20 cases of severe/critical type. Before treatment, the serum phosphorus levels of the two groups were significantly lower than the normal level, and the serum phosphorus levels of the severe/critical patients were lower than those of the common type patients (*t* = 2.767, *P* < 0.010). After treatment, the serum phosphorus levels of the two groups reached normal, and there was no significant difference between the two groups (*t* = 0.231, *P* >0.819). The level of lymphocytes in severe/critical patients was lower than that in normal patients (*t* = 4.636, *P* < 0.001) before treatment. After treatment, the absolute value of lymphocytes in the two groups reached normal, and there was no significant difference between the two groups (*t*=1.208,*P*=0.237). There was a positive correlation between lymphocytes and serum phosphorus, and the correlation coefficient was 0.479.

**Conclusion:** hypophosphatemia is related to the severity of COVID-19, and strengthening the monitoring of serum phosphorus level of COVID-19’s severe/critical patients and correcting hypophosphatemia in time are of significance to improve the prognosis.

## Introduction

The latest threat to global public health is the ongoing outbreak of Coronavirus Disease 2019 (COVID-19). As of March 18, 2020, nearly 210 thousand cases of COVID-19 have been confirmed in more than 100 countries and regions around the world^1^. Phosphorus is one of the essential major elements in the human body. It plays an important role in the phospholipid bilayer that makes up the cell membrane of the body, participates in the synthesis and energy metabolism of adenosine triphosphate (ATP), regulates the enzymatic reaction, and forms a buffer system to maintain acid-base balance in the body^2^. Wei Li et al showed in the study that low phosphorus and chronic kidney disease are the risk factors for the diagnosis of severe pneumonia in the elderly^3^. Liu Bo et al concluded in a meta-analysis of 1555 patients that the level of blood phosphorus can be used as an index to determine the severity of critical illness in critically ill patients^4^.

Since the outbreak of the epidemic, the situation of blood phosphorus in severe/critically ill patients of COVID-19 has not been reported. The purpose of this study was to explore the correlation between hypophosphatemia and the severity of COVID-19 patients by analyzing the level and changes of serum phosphorus in severe /critically ill patients of COVID-19.

## Methods

### Participant

The study enrolled 32 patients from fourth people of Taiyuan City during February 8, 2020 to March 3, 2020 who met the diagnosis and treatment of novel coronavirus pneumonia (trial version fifth)^5^. Informed consent was obtained from all individual participants and the study was approved by Taiyuan Forth People’s Hospital Ethics Committee.

#### Clinical classification standards

According to the novel coronavirus pneumonia treatment plan (trial version fifth), the clinical classification of COVID-19 was: (1) light: mild clinical symptoms, no pneumonia in imaging; (2) common: fever, respiratory tract and imaging findings of pneumonia; (3) heavy: following any of the following: respiratory distress, respiratory rate≥30 Time/min; ② in resting state, the oxygen saturation is≤93%;③ arterial partial pressure of oxygen (PaO2)/concentration of oxygen (FiO2) is≤300 mm Hg (1 mm Hg = 0.133 kPa); (4) severe: one of the following conditions is met: ① respiratory failure occurs and mechanical ventilation is required; ② shock occurs;③ other organ failure needs ICU monitoring and treatment.

#### Determination of serum phosphorus and lymphocyte

Hs-180 automatic biochemical analyzer of Siemens company in Germany was used to determine blood phosphorus by ultraviolet end point colorimetry of phosphomolybdate. The reference value range of blood phosphorus is (0.85-1.51mmol/L). The blood phosphorus <0.85mmol/L was defined as hypophosphatemia.

The original matching detection reagent of Mindray BC-6900 automatic hematology analyzer was used for analysis and determination of blood cells by five classification. The reference range of normal value of lymphocytes is (1.1-3.2×10^9^/L), lymphocytes < 1.1×10^9^/L was defined as low lymphocytes.

#### Grouping and observation index

There were 32 patients diagnosed by COVID-19, including 12 cases of common type and 20 cases of severe / critical type. The general data of the patients were collected according to the case system, including sex, age, epidemiological history, incidence, clinical manifestations, laboratory examination, including nucleic acid test, imaging examination, treatment and outcome, etc. The changes of serum phosphorus level and absolute value of lymphocytes in the two groups were recorded and compared.

#### Statistical method

Mean ± standard deviation was used for statistical description of quantitative data, and *t* test of two independent samples was used for statistical analysis. Qualitative data were described by using rate and composition ratio, and chi square test was used for statistical analysis. Pearson correlation coefficient was used to analyze the correlation between indexes. Statistical significance was set at α=0.05.

## Result

### General information

A total of 32 patients were collected in this study. The average age of severe/critical patients was 49.10±3.93 years old, while that of ordinary patients was 46.75±5.39 years old. In terms of gender composition, the male ratio of severe/critical patients was 60.0%, and the male ratio of common patients was 66.70%. The mean body mass index (BMI) of severe/critical patients was 24.47±2.77, while that of ordinary patients was 24.52±3.33. There was no statistical significance in comparing the general demographic data between the two groups. See Table 1 for details.

**Table 1.** general demographic data of ordinary group and severe/critical group

| variable | severe/critical<br>group | ordinary group | $t/x^2$ | $P$ |
| --- | --- | --- | --- | --- |
| age | 49.10±3.93 | 46.75±5.39 | 0.358 | 0.723 |
| gender |  |  |  |  |
| male | 12 ( 60.0% ) | 8 ( 66.7% ) |  |  |
| female | 8 ( 40.0% ) | 4 ( 33.3% ) | 0.143 | 0.705 |
| BMI | 24.47±2.77 | 24.52±3.33 | 0.043 | 0.966 |

The clinical manifestations of the two groups: 75% of the severe / critical patients had fever and 35% of the patients had dyspnea. In the ordinary group, 58.3% of the patients had fever and 1 case had dyspnea. See Table 2 for details.

**Table 2.** main clinical manifestations of the patients at admission

| groups | fever | cough | dyspnea |
| --- | --- | --- | --- |
| ordinary | 7 ( 58.3% ) | 3 ( 25.0% ) | 1 ( 8.3% ) |
| severe/critical | 15 ( 75.0% ) | 10 ( 50.0% ) | 7 ( 35.0% ) |

#### Comparison of serum phosphorus levels between the two groups before and after treatment

Before treatment, the serum phosphorus level of severe / critical patients was 0.79 ±0.29mmol / L, and that of common patients was 1.11±0.35mmol/L. The level of serum phosphorus in severe / critical patients was lower than that in ordinary patients (*t* = 2.767, *P* < 0.010). After treatment, the serum phosphorus levels of the two groups were normal, and there was no significant difference between the two groups (*t* = 0.231, *P* < 0.819), as shown in Table 3.

**Table 3.** The level of serum phosphorus before and after treatment of two groups

| group | n | before treatment | after treatment |
| --- | --- | --- | --- |
| | | $\bar{x} \pm s$ | $\bar{x} \pm s$ |
| ordinary | 12 | 1.11±0.35 | 1.30±0.42 |
| severe/critical | 20 | 0.79±0.29 | 1.27±0.32 |
| *t* |  | 2.767 | 0.231 |
| *P* |  | 0.010 | 0.819 |

#### Comparison of absolute value of lymphocytes between the two groups before and after treatment

Before treatment, the absolute value of lymphocytes in severe / critical patients was 0.73±0.42 (10^*9^/L), and that in normal patients was 1.45±0.43 (10^*9^/L). The level of lymphocytes in severe / critical patients was lower than that in normal patient)s *t*=4.636,*P*<0.001). After treatment, the absolute value of lymphocytes in the two groups reached normal, and there was no significant difference between the two groups (*t*=1.208,*P*=0.237), as shown in Table 4.

**Table 4.** absolute value of lymphocytes before and after treatment of two groups

| group | n | before treatment | after treatment |
| --- | --- | --- | --- |
| | | $\bar{x} \pm s$ | $\bar{x} \pm s$ |
| ordinary | 12 | 1.45±0.43 | 1.71±0.62 |
| severe/critical | 20 | 0.73±0.42 | 1.42±0.63 |
| *t* |  | 4.636 | 1.208 |
| *P* |  | <0.001 | 0.237 |

#### Correlation analysis between serum phosphorus and absolute value of lymphocytes

There was a positive correlation between the absolute value of lymphocytes and the level of serum phosphorus, and the correlation coefficient was 0.479 (*P* < 0.006), indicating that there was a strong correlation between the two indexes.

## Discussion

Phosphorus is an important element in the body, which participates in the synthesis and energy metabolism of adenosine triphosphate (ATP). The content of total phosphorus in adults is about 700g ^6^, but the concentration of blood phosphorus is only 0.9∼1.6mmol/L ^7^. The clinical incidence of hypophosphatemia is 0.34%-3.10% ^8^. Some studies have found that the incidence of hypophosphatemia in critically ill patients can be as high as 44.8% ^9^. In our study, serum phosphorus levels were lower than normal in 50% of the severe/critical patients, and the level of serum phosphorus in severe / critical patients was lower than that in normal patients. This may be due to the decrease of the level of glycerol 2,3-diphosphate in erythrocytes under the condition of hypophosphatemia, which leads to the left shift of the oxygen dissociation curve, the decrease of oxygen release and the disorder of oxidative phosphorylation process, and finally leads to tissue hypoxia due to the decrease of ATP energy supply^10^. At the same time, the disturbance of lung water absorption is closely related to the metabolic disorder of phosphorus, the lack of phosphorus aggravates the inflammatory level of lung tissue, the ratio of ventilation and blood flow of edematous lung tissue is maladjusted, and the oxygenation function decreases, which can induce fatigue caused by hypoxia in intercostal muscle and diaphragm. At the same time, the production of pulmonary surfactant is reduced, pulmonary alveoli are trapped, and respiratory failure is further aggravated^11^. Some studies have confirmed that timely correction of hypophosphatemia in patients with hypophosphatemia can reverse the above symptoms and improve the prognosis ^12-13^. The level of serum phosphorus is related to the severity of elderly patients with ventilator-associated pneumonia (VAP) and has predictive value for the prognosis of elderly patients with VAP^14-15^. Therefore, the severe damage of immune function leads to hypophosphatemia, which in turn aggravates the progress of the disease.

Lymphocytes are important executors of human immune function. In a healthy state, various lymphocytes and their subsets in the human body interact with each other to maintain the normal immune function of the body. When affected by other factors, such as severe infection or abnormal number or proportion of lymphocytes and their subsets, immune function will be damaged and may lead to a series of diseases ^16^. Data and reports in Hubei Province suggested that the total number of leukocytes in peripheral blood of COVID-19 patients is normal or decreased and lymphocyte count decreases; most patients have a good prognosis after treatment; a few serious patients progress rapidly and peripheral blood lymphocytes decrease progressively. Pathological anatomy of a patient with COVID-19 revealed that the patient’s lymphocytes were over-activated, and the highly pro-inflammatory CCR4 + CCR6 + Th17 cells increased, which partially explained the severe immune damage to the lung of neocoronary pneumonia ^17^. In this study, 85% of patients with severe/critical type had lymphocyte absolute value lower than the reference range, and the lymphocyte level of patients with severe/critical type was lower than that of patients with normal patients. Studies have shown that the immune function of elderly patients with severe pneumonia is significantly lower than that of their healthy peers, while the immune function of patients with hypophosphatemia is lower and generally in the state of immunosuppression. Studies have found that serum phosphorus in elderly patients with severe pneumonia is positively correlated with the ratio of CD4+ / CD8+ ^18^. Our study also showed that there was a positive correlation between the absolute value of lymphocyte and the level of serum phosphorus, so the timely treatment with phosphorus supplementation for severe patients with low phosphorus may improve their immune level and promote their recovery.

The decrease of blood phosphorus level in clinical practice is commonly seen in chronic diseases such as fasting, malnutrition, parenteral nutrition ^19^, metabolic or respiratory alkalosis, diabetic ketoacidosis, alcoholism, chronic alcohol withdrawal, and inadequate phosphorus intake without timely supplementation ^20-21^. Most of the severe/critical COVID-19 patients mentioned in our study were middle-aged, with an overweight BMI and few underlying diseases, and a decrease in blood phosphorus levels was detected 1-10 days after admission. It is more likely that novel coronavirus invades human body, causing a strong stress response and the release of inflammatory factors. On the one hand, gastrointestinal function decline or gastrointestinal mucosal cell integrity and barrier function is impaired, resulting in a reduction in phosphorus intake from food. On the other hand, stress response leads to increased secretion of epinephrine, glucagon, etc., and severely reduced utilization of intracellular phosphorus, leading to the migration of a large amount of phosphorus into the cell to participate in energy metabolism ^22^. In addition to following the treatment of COVID-19 (trial version fifth), we also treated patients with hypophosphatemia with phosphorus supplementation. With the recovery of the disease and the transformation of the viral nucleic acid, the blood phosphorus level and the absolute value of lymphocytes in the majority of severe/critical patients returned to normal. In this process, is it caused by exogenous supplement or redistribution of phosphorus inside and outside cells with the gradual recovery of immune system? More scientific basic or clinical research is needed in the future. In our study, after the patients met the discharge standards of the current COVID-19 treatment scheme (trial version fifth) ^8,^ there were still four patients with absolute value of lymphocytes and two patients with blood phosphorus level not returning to normal. Will these patients have viral nucleic acid conversion to positive again after discharge? There is an urgent need for more clinical research on how to follow up these patients.

In summary, hypophosphatemia was associated with the severity of COVID-19. It is significant to improve the prognosis to strengthen the monitoring of blood phosphorus level in patients with COVID-19 severe/critical type and correct hypophosphatemia in time.

Limitations of this study: 1) the first emergency ward of Taiyuan fourth people’s Hospital is a ward for the treatment of severe/critical COVID 19 patients in Shanxi Province. The small number of samples may affect the statistics and analysis of the results. 2) In the face of sudden outbreaks and life-threatening diseases, no one can use a placebo as a control, so we cannot make sense of exogenous phosphorus supplementation.

## Data Availability

All data used during the study are available.

## Notes

### Competing Interest Statement

The authors have declared no competing interest.

### Funding Statement

The basic applied research projects in Shanxi Province (grant number 201601D021156); The Scientific Research Fund of Shanxi cardiovascular Hospital (grant number 20170203)

